# Postmortem DNA methylation profiling uncovers signatures associated with left ventricle size

**DOI:** 10.1101/2025.08.26.25334499

**Authors:** Steffan Noe Niikanoff Christiansen, Kristine Boisen Olsen, Sara Tangmose Larsen, Pernille Heimdal Holm, Marie Kroman Palsøe, Marie-Louise Kampmann, Stine Bøttcher Jacobsen, Mikkel Meyer Andersen, Jytte Banner, Niels Morling, Jacob Tfelt-Hansen, Jeppe Dyrberg Andersen

## Abstract

1

**Aims:** Postmortem genetic testing is a crucial tool to determine the cause of sudden cardiac death and to enable family testing of relatives. While DNA variants are extensively studied in sudden cardiac death, the role of DNA methylation alterations is poorly understood. In contrast to DNA variants, DNA methylation is tissue-specific. Therefore, it is essential to determine the most relevant anatomical regions for tissue sampling. We aimed to assess the importance of standardised sampling procedures in the investigation of DNA methylation in the human ventricles.

**Methods and results:** We used the Infinium MethylationEPIC array technology to profile the DNA methylation levels among 771,458 CpG sites in the left ventricle, septum, and right ventricle of 15 individuals who died due to sudden cardiac death. We identified 256 differentially methylated regions in the left and right ventricles and the septum. Notably, regions differentially methylated in the left and right ventricles were located in proximity to genes overrepresented in gene ontology terms related to the formation and growth of the human heart. Furthermore, the left ventricle size was correlated with DNA methylation alterations in proximity to the *PITX2* (Pearson’s R^2^ = 0.54, *P* = 0.007) and *PANCR* genes (Pearson’s R^2^ = 0.51, *P* = 0.009). We observed no clustering among the samples from the left and right ventricles and the septum in the examination of all sites. The top 0.1% most variable sites (771 CpG sites) clustered by individual rather than by anatomical origin.

**Conclusion:** We showed that DNA methylation differences of the ventricles near *PITX2* and *PANCR* are associated with left ventricle size. Our findings highlight the importance of standardised tissue collection from multiple regions of the cardiac ventricles in postmortem DNA methylation investigations of the human heart.

## 2 Introduction

Postmortem genetic testing provides valuable and crucial information of the underlying cause of sudden cardiac death (SCD) (Lahrouchi et al. 2017, 2020; Brohus et al. 2021). Beyond the ability to uncover arrhythmogenic disorders that may not be detected through standard autopsy procedures, postmortem genetic testing enables cascade screening of family members. Consequently, postmortem genetics testing is recommended in unexplained SCDs (Stiles et al. 2021; Wilde et al. 2022; Zeppenfeld et al. 2022). Potentially inherited cardiac diseases, including different cardiomyopathies and primary arrhythmogenic disorders of the ventricles, are estimated to account for 43-78% of the SCDs in individuals aged 1-49 years, but in up to 54% of the cases, the cause of death remains unexplained even after autopsy (Lynge et al. 2022). This diagnostic gap underscores need for improved tools to determine the cause of death.

While DNA variants have been extensively studied in postmortem investigations, other molecular alterations that affect the aetiology of SCD remain poorly understood. One promising but underexplored layer of information is DNA methylation. DNA methylation involves the addition of a methyl group to cytosines in CpG dinucleotides and is associated with reduced gene expression in promoter regions (Schultz et al. 2015; Gilsbach et al. 2018). DNA methylation is relatively stable under harsh conditions (Poggiali et al. 2024), while RNA may be more rapidly degraded (Hansen et al. 2014; Jacobsen et al. 2025). This property is crucial when working with samples collected from forensic autopsies where sample quality is highly variable due to tissue decomposition.

DNA methylation has been widely studied in the context of cardiac disease (Haas et al. 2013; Grunert et al. 2016; Meder et al. 2017; Pepin et al. 2019a, 2019b; Sutter et al. 2024; Chen et al. 2025) but its use in postmortem testing has not been well explored. Despite extensive investigations of the role of DNA methylation in cardiac diseases, the heterogeneity of DNA in the human heart is mainly unknown. This gap potentially introduces bias in studies where cardiac tissues are collected from different anatomical locations. To address this, we profiled the DNA methylome of the left ventricle (LV), the septum (S), and the right ventricle (RV). We showed that tissue sampling of different anatomical locations in the ventricles influences DNA methylation. Furthermore, we identified differentially methylated regions (DMRs) among LV, S, and RV located near genes associated with the growth and formation of the chambers of the heart. Last, we identified DMRs between LV and S that were correlated with the volume of the left ventricle.

## 3 Materials and Methods

### 3.1 Design of the study

The heart tissue samples were collected from autopsies of 15 individuals in 2018 at the Section of Forensic Pathology, Department of Forensic Medicine, University of Copenhagen. All deaths were categorised as SCDs. We did not include control samples from individuals that died to other causes than SCD due to challenges of obtaining consent from relatives.

The tissue samples were collected from the mid-ventricular sections of the hearts as part of the standard examination of the heart, as recommended by the Association for European Cardiovascular Pathology (Basso et al. 2017). LV and RV samples were collected from the anterior wall of the ventricles. The samples were only included if they showed no or little decomposition, since high levels of decomposition of cardiac tissues alter the DNA methylation levels (Poggiali et al. 2024). The samples were stored at −70 °C before DNA was extracted for further analyses.

### 3.2 DNA methylation array investigations

DNA from frozen tissue was extracted using the DNeasy Blood & Tissue Kit (Qiagen, Germany) following the manufacturer’s recommendations. The DNA was prepared for methylation measurements with an input amount of 200 ng (Christiansen et al. 2022). The resulting sodium bisulfite-treated DNA was examined using the Infinium MethylationEPIC v2.0 Kit (Illumina, USA) that detects the methylation levels of more than 930,000 positions using a probe-based approach.

### 3.3 Data analysis

The data analysis was performed in the statistical environment R (version 4.3.1) using the *tidyverse* (version 2.0.0) (Wickham 2014; Wickham et al. 2019), and *patchwork* (version 1.2.0) (Pedersen 2024) packages.

The *SeSAMe* package (v1.20.0) was used to convert the raw intensity (.idat) data files into normalised methylation values using the *openSesame* function with the default preprocessing code (“QCDPB”) (Zhou et al. 2018). In brief, the preprocessing steps include masking poorly designed probes (“Q”), inferring channels for Infinium I probes (“C”), performing non-linear dye bias correction (“D”), removing probes with high out-of-band signals (“P”), and conducting background subtraction using out-of-band signals (“B”). The probes were annotated with the “EPICv2.hg38.manifest.gencode.v41.tsv” annotation file obtained from https://github.com/zhou-lab/InfiniumAnnotation (Kaur et al. 2023). Only data from chromosomes 1-22 were included in the analyses.

Methylation levels from array experiments were reported as either β-values or M-values (Supplementary Methods 1). In brief, the β-values were calculated as the ratios of methylated signal compared to the total signal (reported as a value between 0 and 1). The M-values were reported as values between -Infinity and Infinity. The M-values were preferred for analyses because the variance is approximately constant (Du et al. 2010). To address unwanted batch variation in the normalised methylation values, the *ComBat* function from the *sva* package (version 3.48.0) (Leek et al. 2012) was used (Supplementary Methods 2). The adjusted M-values were used for the principal component analysis (PCA) and the exploration of the most variable positions. The PCA was conducted using the base R *prcomp* function in R with centred and scaled values. The four components explaining the largest fraction of variability were visualised. Heat maps were visualised with the *pheatmap* package (version 1.0.12) with the Euclidean distance as a similarity measure and complete linkage for clustering. Pearson’s correlation coefficient was calculated to estimate the correlation among the samples. The squared correlation coefficient (*R*^2^) was reported since this value is equal to the fraction of systematic variation between two variables.

### 3.4 Differential methylation

Differentially methylated loci were identified in three pairwise comparisons (LV versus RV, LV versus S, and RV versus S) using the *DML* function from the *SeSAME* package (v1.20.0) (Zhou et al. 2018) with the following input formula: (β ∼ origin + id), where origin refers to the tissue section (LV, S or RV) and id refers to the tissue donor id. The output from the *DML* function was used to identify differentially methylated regions (DMR) using the *DMR* function from the *SeSAME* package (Zhou et al. 2018). In brief, the differentially methylated positions were combined in regions based on Euclidean distances. Next, a combined *p*-value was calculated using Stouffer’s method (Stouffer et al. 1949). The resulting *p*-values were adjusted with the *p.adjust(method = “BH”)* function in R. DMRs with a minimum of 5 CpGs and an adjusted *p*-value below 0.05 were considered significant and used for further analyses. The DMRs were annotated to nearby genes based on the *EPICv2.hg38.manifest.gencode.v41.tsv* manifest file (Kaur et al. 2023), available from https://zwdzwd.github.io/InfiniumAnnotation. Ensembl gene names were converted to gene names using the *getBM* function from the *biomaRt* R package (version 2.56.1) (Durinck et al. 2009). Overlapping DMRs were visualised with the *ggVennDiagram* R package (version 1.5.2) (Gao et al. 2024) and nearby genes were visualised with the *ggtranscript* R package (version 0.99.9) (Gustavsson et al. 2022).

### 3.5 Enrichment analysis

Gene ontology (GO) term overrepresentation analysis was conducted with the *clusterProfiler* R package (version 4.8.3). The *enrichGO* function was used to test the association between genes located in proximity to the DMRs and the two terms, “biological processes” and “molecular function”. The *org.Hs.eg.db* (version 3.17.0) annotation package was used as the input database. Genes with one or more annotated DMRs were included in the background set as input for the *enrichGO* function. The *p*-values were adjusted with the default method (*pAdjustMethod = “BH”*) of the *enrichGO* function. Adjusted *p*-values below 0.05 were considered statistically significant. Overrepresentation analyses can be biased if a DMR is annotated to multiple nearby genes that share the same gene ontology (GO) term. Therefore, we excluded a single gene ontology term (GO:0070003), which is driven only by a single DMR near two genes (*PSMB8* and *PSMB9*).

### 3.6 Postmortem computed tomography scans

Before the autopsy, the deceased individuals underwent a whole-body unenhanced computed tomography (CT) scan as part of the routine investigation. These postmortem CT scans were performed on a Siemens Somatom Definition 64 slice scanner (Siemens Healthcare, Erlangen, Germany) using a standard protocol with a tube voltage of 120 kV and automatic tube current modulation (reference mAs: 300 and effective mAs range: 240-300). Images of the thorax were reconstructed with 2.5 mm slice thickness using a standard soft tissue kernel. In one case, only the bone kernel reconstruction was available.

The LV and RV volumes (in cm^3^) were estimated from the CT images that were assessed using Myrian Software version 2.12.3 (Intrasense, Montpellier, France). The LV and RV volumes were calculated by manually contouring the RV and LV, using the “filled space” function of the region of interest (ROI) on each axial slice, beginning from the demarcation of the ventricles from the atria to the apex. No interpolation between slices was applied. The images were viewed using a soft tissue window with a window level of 40 HU and a window width of 340 HU (−135 HU to +215 HU), and no smoothing or automatic segmentation tools were used. Each volume included the ventricular wall and the associated cavity, as it was not possible to separate them due to the lack of attenuation differences between the cavity and the surrounding ventricular muscle in the images. The epicardial fat was excluded. The interventricular septum was included in the LV volume. Image analysis was performed by a forensic pathologist with postmortem CT certification and more than 15 years of experience.

### 3.7 Ethics approval and consent to participate

Ventricular tissue was collected from the research biobank at the Section of Forensic Pathology, the Department of Forensic Medicine, University of Copenhagen (004-0064/21-7000 and 004-0022/18-7000). The study was conducted according to the Declaration of Helsinki and is registered at the University of Copenhagen’s joint records of processing of personal data in research projects and biobanks (KU j.nr 514-0726/22-3000). The project complies with the rules of the General Data Protection Regulation (Regulation (EU) 2016/679). The study was approved by the Committees on Health Research Ethics in the Capital Region of Denmark (H18014656). The relatives of the deceased individuals received written information about the project and informed consent was obtained and documented by physicians according to Danish legislation (“VEJ nr 9267 af 13/06/2013” issued by The Ministry of the Interior and Health).

## 4 Results

### 4.1 DNA methylation of the ventricles of the human heart

Cardiac tissue was collected from 15 deceased individuals (13 males and two females) to investigate the DNA methylation patterns of the ventricles of the human heart (Figure 1A). The samples were collected from individuals who died due to SCD. Tissue was collected from LV, S, and RV from the midventricular section. The boundaries of the age intervals ranged from 25-50 years (median 46 years).

**Figure 1.**
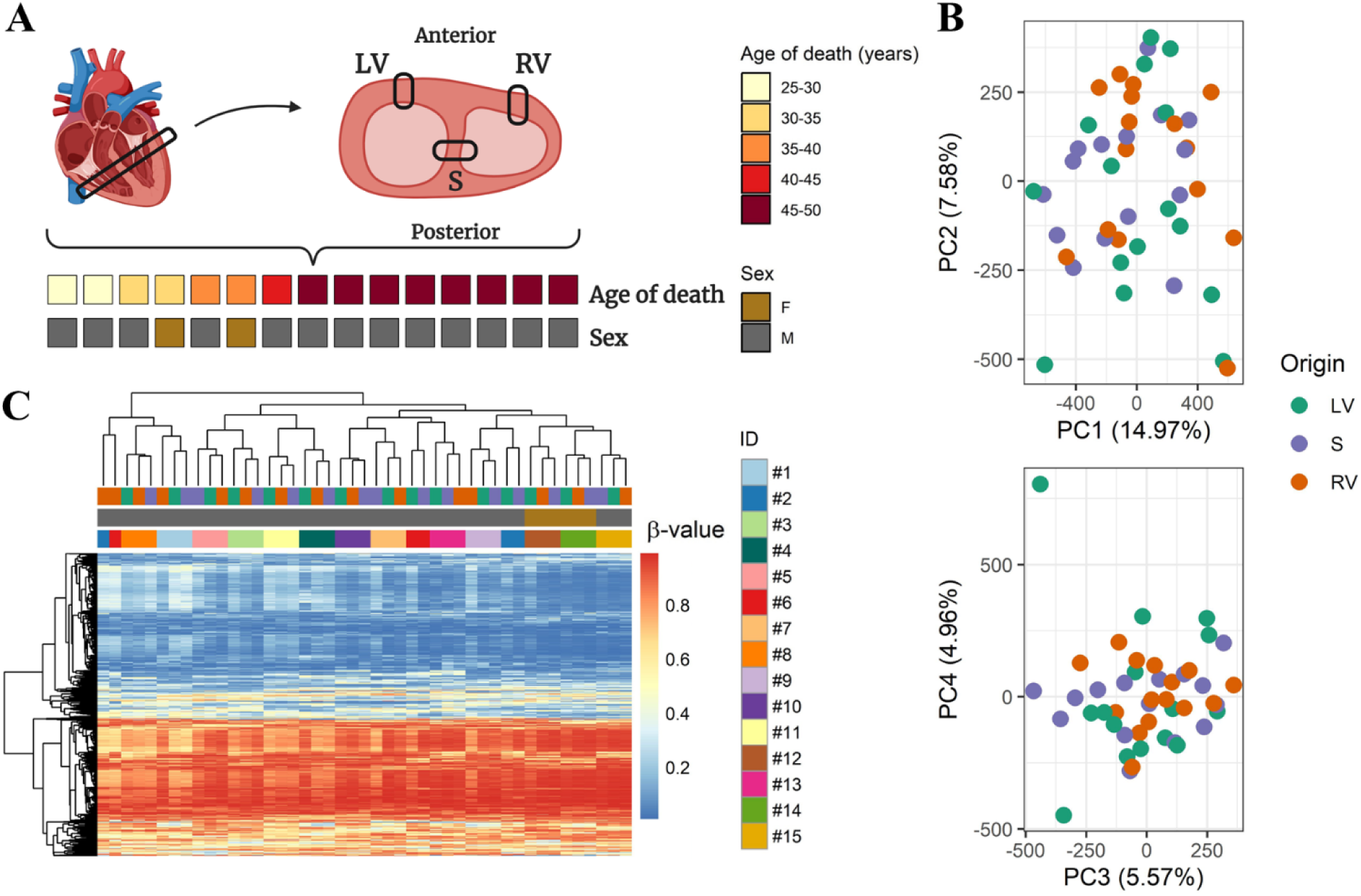
DNA methylation patterns of the ventricles of the heart. (A) Overview of the study population characteristics and the sampling procedure from the left ventricle (LV), septum (S), and the right ventricle (RV). (B) Principal component (PC) analysis of M-values from 771,458 positions coloured according to the origin of the tissue samples. (C) Heatmap of the 0.1 percentile of the positions (771 CpG positions) with the highest variability among the samples. The colours below the dendrogram represent the origin of the samples and sex according to (A) and (B).

In the first step of the analysis, the data were normalised and filtered to omit data from CpG sites with low-intensity signals. We only included data from CpG sites annotated to the autosomal chromosomes (chromosomes 1-22). After this step, data from 771,458 CpG sites remained. The 65 identification SNPs of the Infinium MethylationEPIC v2.0 array were used to exclude possible mislabelling of the paired samples from each individual (Fig. S1).

All samples showed the characteristic bimodal β-value distribution, which is expected for DNA methylation values from array analysis (Fig. S2). The squared Pearson’s correlation coefficients (R^2^) of the β-values ranged from 0.961 to 0.995 (median: 0.987) among all samples. In comparison, the median values of R^2^ between LV, S, and RV of each individual ranged from 0.978 to 0.994 (median of medians: 0.991) (Fig. S3).

The overall variability of the data was investigated with a PCA of M-values (logit-transformed β-values). The first four components explained 33.1% of the total variation (Figure 1B). We observed no overall tendency for clustering based on the variability of all sites. Next, we investigated the variability among the 0.1% most variable positions corresponding to 771 CpG positions (Figure 1C). The inter-individual variability of the samples exceeded the intra-individual variability in all samples except for those obtained from two individuals (ID #2 and #6), as shown by the dendrogram above the heatmap (Figure 1C), where RV deviated from LV and S in both individuals.

### 4.2 Differential methylation among the left and right ventricles, and the septum

The analysis of differential methylation was separated into three pairwise comparisons (LV vs RV, LV vs S, and RV vs S). A DMR refers to a specific genomic region where the DNA methylation levels are different in at least one of the pairwise comparisons (Figure 2A). In the three pairwise comparisons, 64 (LV-RV), 40 (LV-S), and 186 (RV-S) significant DMRs with 5 ≥ CpG positions were identified (Figure 2B and Table S1). However, since multiple DMRs were observed in two different pairwise comparisons, the total number of DMRs was 256, corresponding to the sum of the values within the Venn diagram.

**Figure 2.**
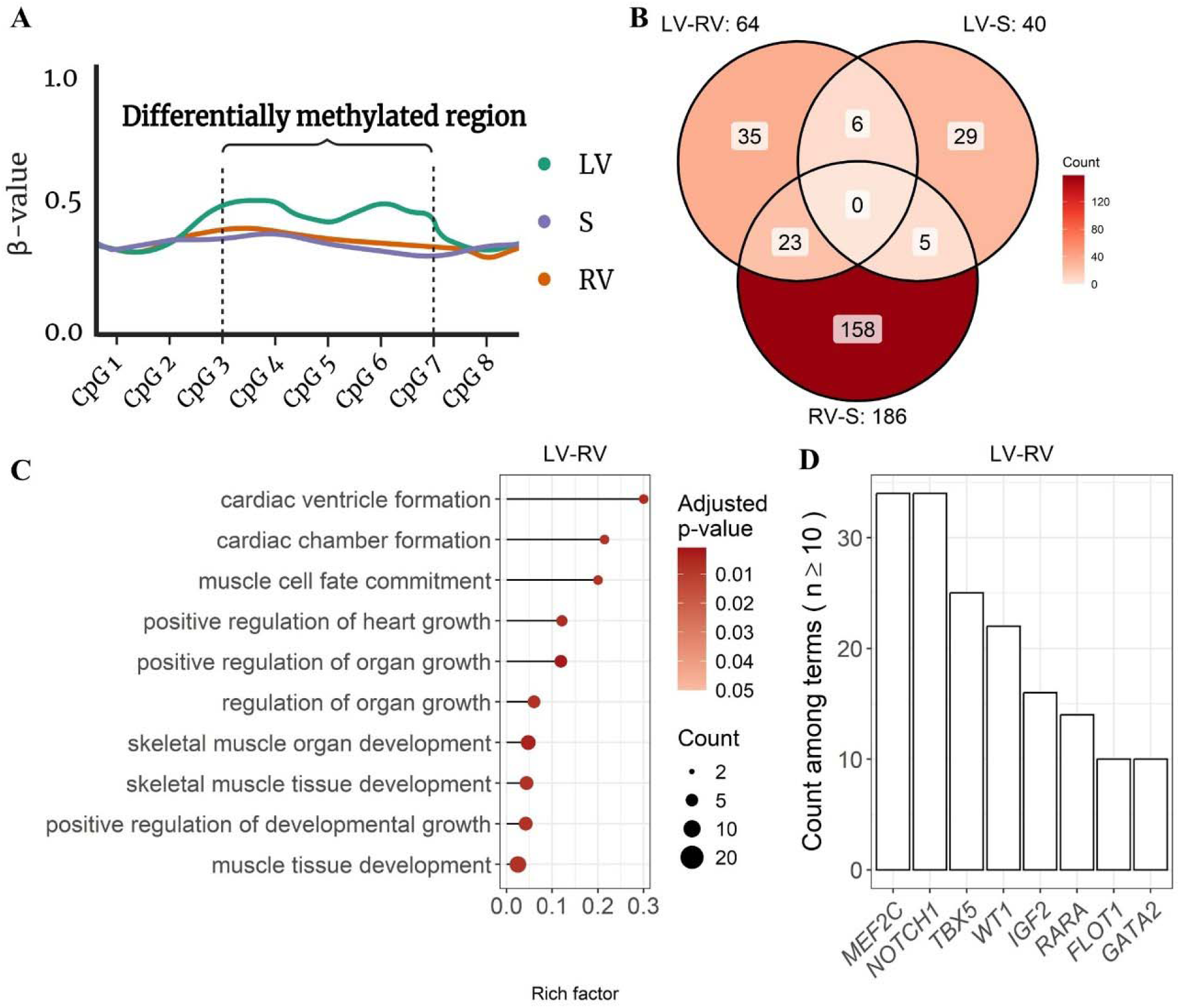
Differentially methylated regions of the ventricles. (A) Illustration of a differentially methylated region (DMR) with 5 CpG sites. The DMRs were investigated in three pairwise investigations among the left ventricle (LV), septum (S), and right ventricle (RV). In this illustration, the region was differentially methylated between LV and S and between LV and RV, illustrating the potential overlap among the identified DMRs in the three pairwise investigations. (B) Counts of differentially methylated regions (DMR) among the three pairwise comparisons (LV-RV, LV-S, and RV-S). (C) Gene ontology (GO) overrepresentation analysis of biological processes among the DMRs between LV and RV. The ten terms with the highest rich factor (ratio of DMR-genes divided by all genes in the specific term) are visualised. (D) Count of Gene Ontology terms that each gene is assigned to (genes associated with < 10 significant terms are not included in the figure).

The mean β-value differences of the DMRs ranged from −0.05 to 0.05 (LV-RV), from −0.046 to 0.062 (LV-S), and from −0.066 to 0.041 (RV-S) (Table S1). A positive mean β-value difference of the LV-RV comparison indicated that the DNA obtained from LV samples had higher methylation levels than samples from RV. In contrast, a negative value corresponded to a DMR with lower methylation levels in LV compared to RV. The majority of the 256 DMRs were detected in only one of the three pairwise comparisons (Figure 2B). However, 34 DMRs (corresponding to the sum of 23, 6, and 5 overlapping DMRs) were found in two of the three pairwise comparisons. In these cases, one tissue, e.g. LV, was differentially methylated relative to both S and RV, as illustrated in Fig. 2A. The median width among the 256 DMRs was 2,130 bp (interquartile range: 794-11,272 bp) and included information on DNA methylation levels of up to 30 CpG positions.

The DMRs were assigned to nearby genes (DMR-genes) to examine the potential biological role of the DMRs. Among the DMRs, 47 could not be assigned to any genes, while the remaining DMRs were assigned to between one and five genes. We conducted an overrepresentation analysis using the *clusterProfiler* R package (Wu et al. 2021). Overrepresentation analysis is a method that can be used to identify whether certain GO-terms of “*Biological Processes*” or “*Molecular Functions*” are more common among a set of genes than expected by chance.

The genes from the three comparisons (LV-S, LV-RV, and RV-S) were overrepresented among 46 biological processes after correction for multiple testing (Table S2), showing that these genes were observed more often than expected in each of these biological processes. Among the total of 46 overrepresented biological processes, 45 were observed for the LV-RV comparison (Figure 2C). The DMR genes of this comparison were overrepresented in terms such as “*cardiac chamber formation*”, “*cardiac ventricle formation*”, and “*positive regulation of heart growth*”. Of these, the three most frequently involved genes were *MEF2C*, *NOTCH1*, and *TBX5* (Figure 2D). There were no overrepresented biological processes of the genes for the LV-S comparison. One biological process (“*cell morphogenesis involved in neuron differentiation*”) was statistically significantly enriched for the RV-S comparison.

The DMR-genes were also overrepresented among seven molecular function terms for the LV-RV comparison and two terms for the RV-S comparison (Table S3). The overrepresented molecular functions included “*DNA-binding transcription activator activity*”, *“transcription coregulator binding*”, and “*transcription coactivator binding*”.

### 4.3 Molecular signatures of *PITX2* associated with left-right sidedness

Two DMRs overlapped either “paired-like homeodomain transcription factor 2” (*PITX2*) or “*PITX2* adjacent noncoding RNA” (*PANCR*) (Figures 3A-B). The first DMR, hereafter labelled as the *PANCR* DMR, overlapped *PANCR* at position chr4:110,610,770-110,612,796 and was identified in the LV-RV as well as the LV-S comparison (Figure 3A, left panel, and Figure 3B). The other DMR, hereafter labelled as the *PITX2* DMR, was located at position chr4:110,615,611-110,618,136 overlapping the 3’ untranslated region of *PITX2* and was only identified in the LV-S comparison (Figure 3A, right panel, and Figure 3B).

**Figure 3.**
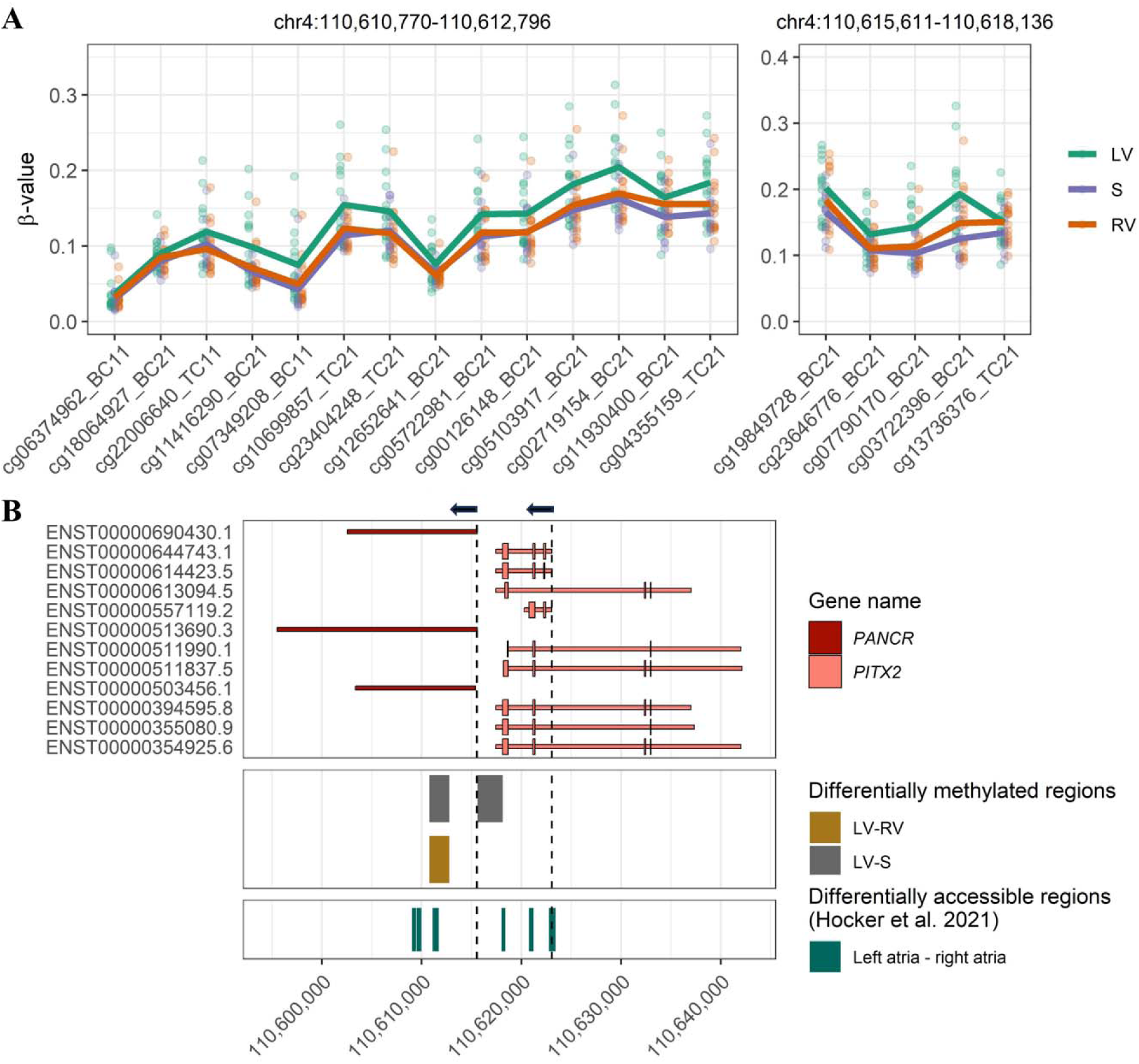
Differentially methylated regions at the *PANCR-PITX2* locus. (A) β-values per position are shown for two differentially methylated regions (DMRs) located in proximity to “paired-like homeodomain transcription factor 2” (*PITX2*) and “*PITX2* adjacent noncoding RNA” (*PANCR*). The mean β-values per position among each tissue are shown as lines covering the positions. Each value on the horizontal axis corresponds to the unique identifier of the positions with differential methylation. (B) Position and direction of transcription for *PANCR* and *PITX2*. The *PANCR* DMR (chr4:110,610,770-110,612,796) was differentially methylated in two comparisons (LV-RV and LV-S) while the *PITX2* DMR (chr4:110,615,611-110,618,136) was differentially methylated only in the comparison of LV-S. Both DMRs overlapped with previously identified differentially accessible regions between left and right atria (Hocker et al. 2021). ENST00000644743.1 correspond to the *PITX2c* isoform. The vertical dashed lines indicate the location on previously identified transcribed regulatory elements between the left and right side of the heart (Deviatiiarov et al. 2023). Due to their proximity to each other, the two transcribed regulatory elements associated with *PANCR* cannot be distinguished in this figure. The arrows indicate the orientation of transcription. LV: Left ventricle, S: septum, RV: right ventricle.

To examine the transcriptional regulatory role of these DMRs, we compared our findings to previously published single-nuclei chromatin accessibility data (Hocker et al. 2021) and transcribed regulatory elements (Deviatiiarov et al. 2023) in samples from the left and right sides of the human heart. Single-nuclei profiling of chromatin accessibility is a method for identification of active regulatory regions that control gene expression in specific cell types (Cusanovich et al. 2015). We observed no differentially accessible regions between LV and RV that overlapped the *PANCR* DMR and the *PITX2* DMR. However, two differentially accessible regions between left and right atria overlapped both the *PANCR* and *PITX2* DMRs, and four other differentially accessible regions overlapped either the *PANCR* or the *PITX2* DMR (Figure 3B). Notably, all six differentially accessible regions were specific for cardiomyocytes and were more accessible in the left than the right atrium.

We compared the location of the DMRs to transcribed regulatory elements based on cap analysis of gene expression of samples from the left and right sides of human hearts (Deviatiiarov et al. 2023). Among 35 transcribed regulatory elements that were more active in the left compared to right side of the heart, three were annotated to *PANCR* or *PITX2* (Fig. 3B and Fig. S4). One transcribed regulatory element was located at the 5’ end of the ENST00000644743.1 transcript corresponding to the *PITX2c* isoform and located 4,933 nt away from the *PITX2* DMR. The two other transcribed regulatory elements were located only 24 nt away from each other and 33 nt away from the *PITX2* DMR.

### 4.4 Correlation of *PITX2* locus differential methylation with left ventricle volume

Due to the previous observations of differential accessibility and transcriptional activity, combined with our observation of the *PANCR* and *PITX2* DMRs, we proceeded to compare the β-values of the DMRs with the volume of LV as measured from postmortem CT scans.

There were no available CT measurements from three of the individuals. In two cases, postmortem CT was not performed because the scanner was unavailable at the time of the autopsy (#10 and #11). A third case (#14) was excluded due to a large amount of right-sided intraventricular air.

As expected, the heart weight was positively correlated with the volume of LV and S obtained from the CT scans (Pearson’s R^2^ = 0.94, *P* = 1.57×10^-7^) (Fig. S5). To account for the expected correlation between LV volume and body size, we adjusted the LV volume for body surface area. We compared the measure to the median β-value difference for the LV-RV and LV-S comparisons of the *PANCR* DMR and the LV-S comparison of the *PITX2* DMR (Figure 4). There was no significant correlation for the LV-RV comparison (*P* = 0.341). However, the median β-value differences were negatively correlated with the LV volume for the *PANCR* DMR (Pearson’s R^2^ = 0.51, *P* = 0.009) and the *PITX2* DMR (Pearson’s R^2^ = 0.54, *P* = 0.007) in the LV-S comparison. In other words, reduced DNA methylation of LV compared to S was associated with large LV volumes.

**Figure 4.**
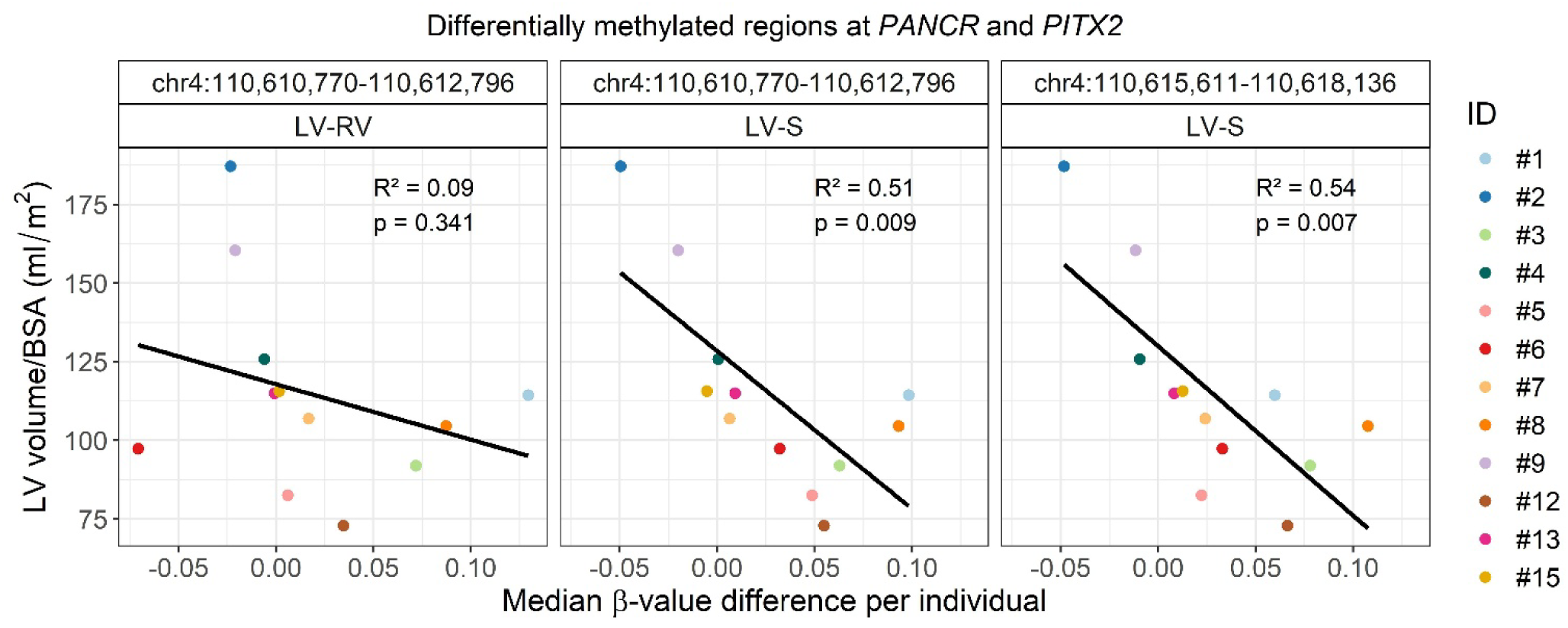
Association of *PANCR-PITX2* DNA methylation with left ventricular volume. Left ventricle volume corrected for body surface area (BSA) against the median β-value difference of each individual within the differentially methylated region assigned to *PANCR* and *PITX2*. Pearson’s squared correlation coefficient (R^2^) and *p*-values are shown in the upper right corner of the panels. The black lines indicate the regression lines based on a linear model.

## 5 Discussion

### 5.1 Standardised tissue collection for postmortem investigations

In multiple studies, DNA methylation alterations of the human heart have been linked to different cardiac diseases (Haas et al. 2013; Grunert et al. 2016; Meder et al. 2017; Pepin et al. 2019a, 2019b; Sutter et al. 2024; Chen et al. 2025). However, a significant challenge in many studies is the inability to consistently collect tissue from the same anatomical locations of the heart. Therefore, we standardised cardiac tissue collection in forensic autopsies and identified considerable differences between tissue sections from the human heart.

Consistent with a previous investigation of DNA methylation from LV and RV from two individuals (Schultz et al. 2015), we found that the global DNA methylation signatures of cardiac ventricle samples are more similar within individuals than among individuals (Figures 1B-C). Despite this similarity, we identified DMRs among LV, S, and RV and showed their association with critical processes such as development, growth, and formation of the human heart (Figures 2C-D). In addition, the DMRs are assigned to well-known disease-associated genes. For instance, *TBX5* and *NOTCH1* are considered definitive genes for congenital heart disease (Morton et al. 2022), while common variants assigned to *TBX5* and *PITX2* are associated with atrial fibrillation (Christophersen et al. 2017; Roselli et al. 2018, 2025). Based on our observations of specific DNA methylation signatures of distinct anatomical locations, we highlight the necessity of examining samples in a standardised manner to avoid spurious findings in DNA methylation investigation of cardiac diseases.

### 5.2 Differential methylation of *PANCR* and *PITX2*

We observed two DMRs in proximity to *PITX2* and *PANCR* (Figure 3A) in line with the previous findings that *PITX2* and *PANCR* are differentially expressed between the left and the right atrium (Gore-Panter et al. 2016; Cranley et al. 2024). *PITX2* promoter methylation was previously associated with reduced *PITX2* gene expression in human induced pluripotent stem cells (Madsen et al. 2020) and reduced Pitx2 protein levels in mice (Chen et al. 2025). Therefore, to evaluate the regulatory potential of the DMRs, we compared them to differentially accessible regions and transcribed regulatory elements between left and right sides of the heart (Figure 3B). As expected, both features overlapped the assumed promoter region of the *PITX2c* isoform known for being specifically expressed in left atria compared to right atria (Kirchhof et al. 2011). In contrast, the two DMRs did not overlap the promoter region of *PITX2*. However, the *PANCR* DMR overlapped a differentially accessible region while the *PITX2* DMR was located less than 33 and 71 nt upstream the two transcribed regulatory elements of *PANCR*. Since *PITX2* expression is regulated by *PANCR* (Gore-Panter et al. 2016), it is possible that the observed DNA methylation patterns reflect an indirect mechanism for regulation of *PITX2* expression.

*PITX2* and *PANCR* are expressed considerably less in the adult ventricles than in adult left atria (Bayraktar et al. 2024; Cranley et al. 2024). Therefore, it may seem surprising that we observe DMRs in adult ventricles where *PITX2* is typically expressed in barely detectable levels. However, nuclear *Pitx2* is induced in adult ventricles of mice after myocardial infarction (Tao et al. 2016). Thus, the observed DNA methylation alterations may be a response to undetected heart injury. Alternatively, the *PITX2* and *PANCR* DMRs may be a stable signature from the past. Less than 1% of cardiomyocytes are replaced in the adult human heart each year (Bergmann et al. 2015). Therefore, DNA replication-dependent removal of DNA methylation signatures by passive demethylation should be considerably lower in cardiac tissue than in tissues with a high turnover.

### 5.3 Left ventricle volume associated with differential methylation of *PANCR* and *PITX2*

Recently, it was shown in a genome-wide association study that genetic variants assigned to *PITX2* were associated with the BSA-adjusted volume of left atria (Pirruccello et al. 2024). The identified lead variant (rs2634073) was located at position chr4:110,744,627, more than 120kb away from the *PITX2* gene. Here, we demonstrated that DNA methylation differences between LV and S in both the *PITX2* and *PANCR* DMRs were associated with LV volume as measured by postmortem CT scanning (Fig. 4). Consistent with these two findings, it was shown that atrial-specific *Pitx2* reduction enlarged the size of both the atria and ventricles in a murine model (Chinchilla et al. 2011). Interestingly, during the embryological state, morphological alterations were only detected in the atria, but not in the ventricles, suggesting that the ventricular alterations are likely secondary to the dysfunction of the atria.

### 5.4 Strengths and limitations of the study

While CT-based ventricular volumetry is an established method in living subjects (Goo and Park 2015), a similar standardised image acquisition is not possible in autopsy cases. Unlike in living patients, the use of contrast agents and the ability to perform imaging during specific phases of the cardiac cycle, such as end-diastole, are not feasible postmortem. Hence, varying degrees of blood pooling in the ventricles can be observed postmortem, and the use of non-enhanced postmortem CT further complicates the differentiation of anatomical structures due to the lack of contrast between tissues.

Almost half of the included cases exhibited mild decomposition at the time of autopsy, which poses a risk of cardiovascular gas accumulating within the heart chambers, potentially leading to overestimation of ventricular volumes (Jackowski et al. 2006). In one case (#2), a hemopericardium, i.e. the presence of blood in the pericardial sac, caused cardiac compression, resulting in reduced ventricular volumes. In another case (#9), a surgical implant in the thoracic wall caused imaging artefacts, which may have affected the measurements. Image analysis was performed by a single reader without assessment of intra- or inter-observer variability. Therefore, some degree of subjectivity and variability in the manual contouring of the images must be anticipated. Despite these limitations, the estimated ventricular volumes showed, as expected, a strong correlation with heart weight.

We conducted, to our knowledge, the most comprehensive characterisation of DNA methylation in the human ventricles to date. However, we acknowledge the limitations of the unbalanced sex distribution of our study. Male individuals are overrepresented in forensic autopsies (Larsen and Lynnerup 2011) and among autopsied SCDs (Winkel et al. 2017), which is reflected in this study. In the data analysis, we filtered the DMRs for the number of CpG sites and significance levels. However, we did not filter for effect size for two reasons. First, since the analysis was conducted on bulk tissue, a stringent effect size threshold would possibly exclude DMRs only present in a single cell type, e.g. cardiomyocytes. Second, due to the heterogeneity of the cause of death among young individuals, we expected heterogeneous DNA methylation patterns at genomic regions associated with any underlying disease causing SCD. While this decision possibly resulted in DMRs driven by high effect sizes in only a subset of the individuals, we demonstrated that the function of the identified DMRs and nearby genes was primarily related to biological processes necessary for cardiac and muscle growth and development.

## 6 Conclusion

Our findings showed that DNA methylation alterations near *PITX2* and *PANCR* are associated with left ventricle size. The study demonstrates the importance of collecting samples from multiple regions of the cardiac ventricles in DNA methylation investigations of the human heart.

## Data Availability

The data are available from the corresponding authors upon reasonable request.

https://zenodo.org/records/17406594

## 8 Acknowledgements

We thank Anja Ladegaard Jørgensen for technical assistance with this work. We sincerely thank all the relatives for their consent to this study.

## 10 Author contributions

S.N.N.C., M.L.K., J.B., N.M., J.T.H., and J.D.A. designed the study. K.B., S.T.L., P.H.H., M.K.P., and J.B. conducted the autopsies and postmortem CT scans, collected the tissue, and obtained informed consent for the study from the relatives of the deceased individuals. S.B.J. and M.L.K. processed the samples and conducted the experiments. S.N.N.C., K.B., S.T.L., M.M.A., and J.D.A. performed the data analysis. S.N.N.C., K.B., S.T.L., J.T.H., and J.D.A. wrote the initial draft of the manuscript. All authors revised and approved the final manuscript.

## 11 Abbreviations

CT: Computed tomography
DMR: Differentially methylated region
GO: Gene ontology
LV: Left ventricle
*PANCR*: *PITX2* adjacent noncoding RNA
PC: Principal component
PCA: Principal component analysis
ROI: Region of interest
RV: Right ventricle S: Septum
SCD: Sudden cardiac death

## 12 Availability of data and materials

The data are available from the corresponding authors upon reasonable request. The “EPICv2.hg38.manifest.gencode.v41.tsv” annotation file was obtained from https://github.com/zhou-lab/InfiniumAnnotation. External data used for this study are available from the online versions of the manuscripts. The transcribed regulatory elements were available from the online version of the paper published in Nature Cardiovascular Research (https://doi.org/10.1038/s44161-022-00182-x). The differentially accessible regions were available from the online version of the paper published in Science Advances (https://www.science.org/doi/10.1126/sciadv.abf1444).

The code for the analyses and the figures is available from GitHub (https://github.com/SteffanChristiansen/DNA_methylation_ventricles) and Zenodo (https://doi.org/10.5281/zenodo.17406230). Mean β-values of left and right ventricle and septum tissue among all sites is available from Zenodo (https://zenodo.org/records/17406594).

