## Supplementary methods 1-2 and supplementary figures 1-5 for "Postmortem DNA methylation profiling uncovers signatures associated with left ventricle size"

**Supplementary material**

### Supplementary methods

#### Supplementary methods 1

**Methylation values**

Methylation levels from array experiments were reported as either β-values or M-values. The β-values are calculated as the ratios of methylated signal compared to total signal (reported as a value between 0 and 1). The β-values were calculated as indicated below.

$$\beta=\frac{methylated signal}{unmethylated signal+methylated signal}$$

M-values (reported as a value between -Infinity and Infinity) are preferred for analyses where it is assumed that the variance are approximately constant (Du et al. 2010). The relationship between β-values and M-values is shown below.

$$M={log}_{2}\left( \frac{\beta}{1-\beta} \right); \beta= \frac{2^{M}}{2^{M}+1}$$

#### Supplementary methods 2

**Batch correction**

To address unwanted batch variation in the normalised methylation values, the *ComBat()* function from the *sva* package (Leek et al. 2012) was used. In brief, this approach is based on a Bayes framework and enables adjustment for known covariates (Johnson et al. 2007). The batch correction was conducted with M-values and default parameters for adjustment of known batches. The function was applied in stages: first to adjust batch effects from the slides, and then to correct for the position of each array on the slide. Adjusted β-values were calculated with the *MValueToBetaValue()* function from the *SeSAMe* package (Zhou et al. 2018) and were used for the remaining analyses and visualisation.

### Supplementary figures

#### Supplementary Figure 1

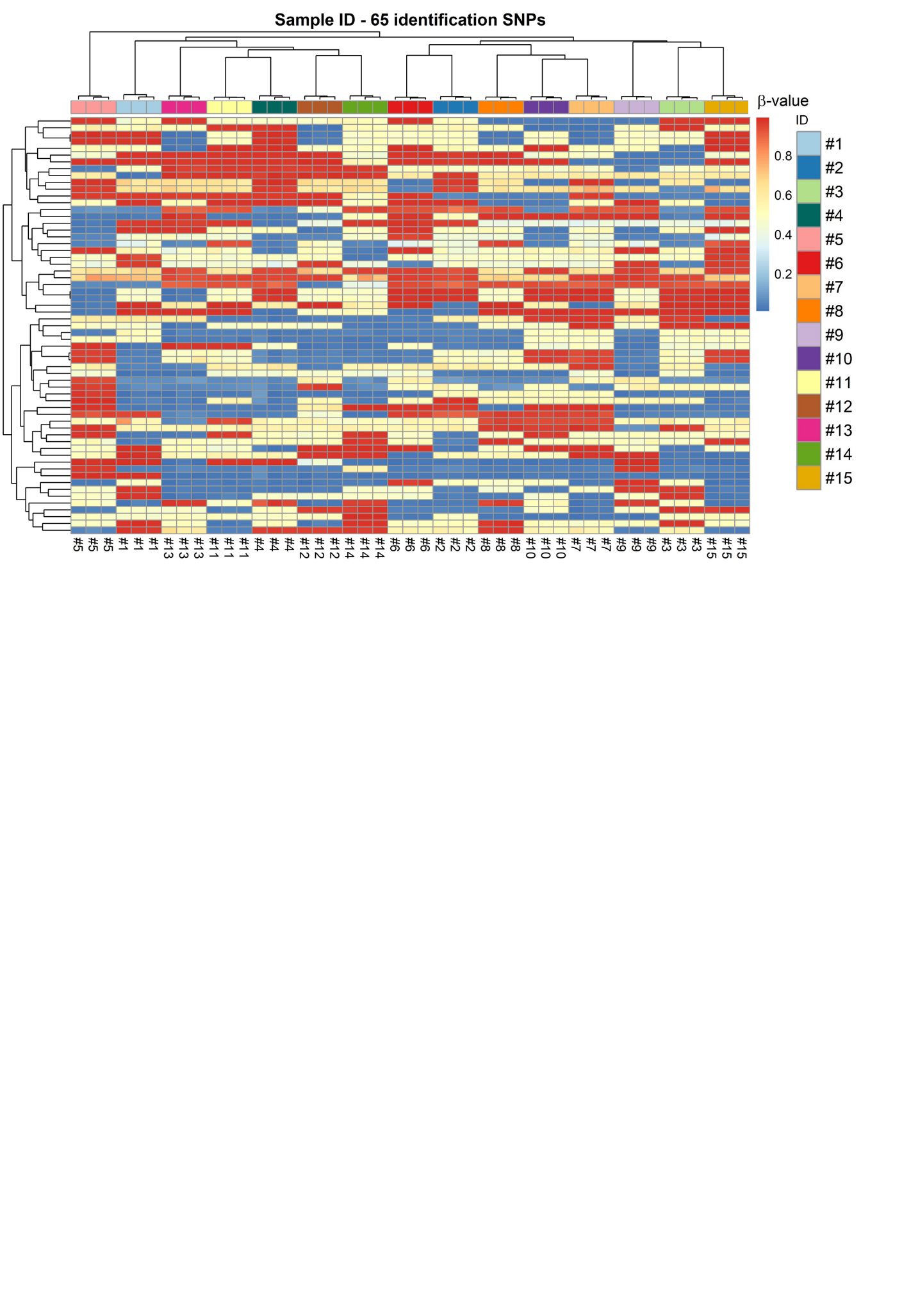

**Sample identification using the identification single nucleotide polymorphisms.** Two-dimensional dendrogram with a heatmap of the β-values of the single nucleotide polymorphism (SNP) typing results of three samples from each of included individuals. Each row of the heatmap represents an SNP, and each column represents a sample. The colours and their intensities illustrate the β-values of the 65 identification SNPs. The β-values generally grouped into three clusters around 0, 0.5, and 1 corresponding to homozygous, heterozygous, and opposite homozygous SNP typing results, respectively.

#### Supplementary Figure 2

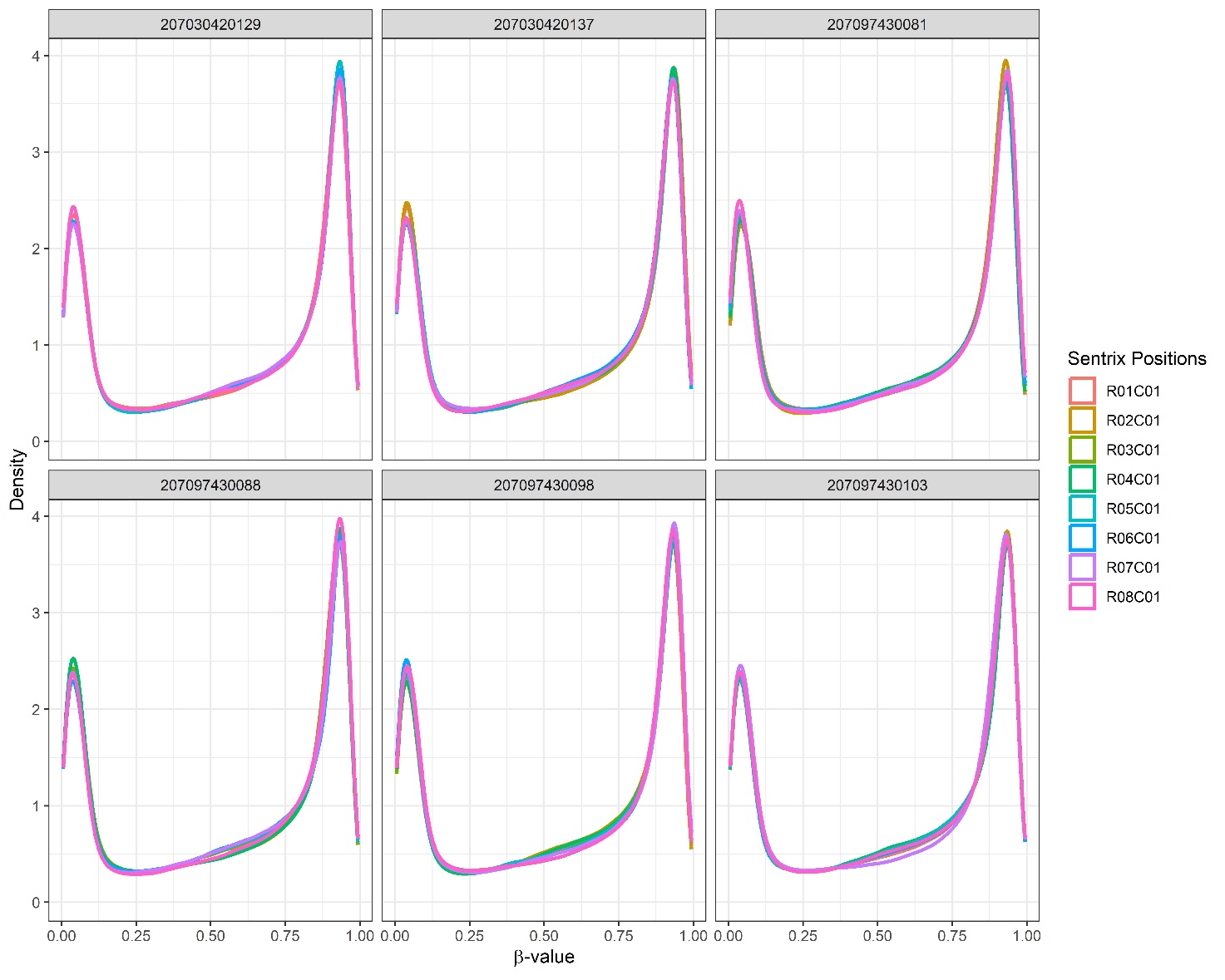

**β-value distributions.** Density plots of β-values from 771,458 positions investigated on six different slides and eight different positions on the slides (sentrix_position). The density estimates were based on a Gaussian kernel using the *geom_density* function within the *ggplot* R package.

#### Supplementary Figure 3

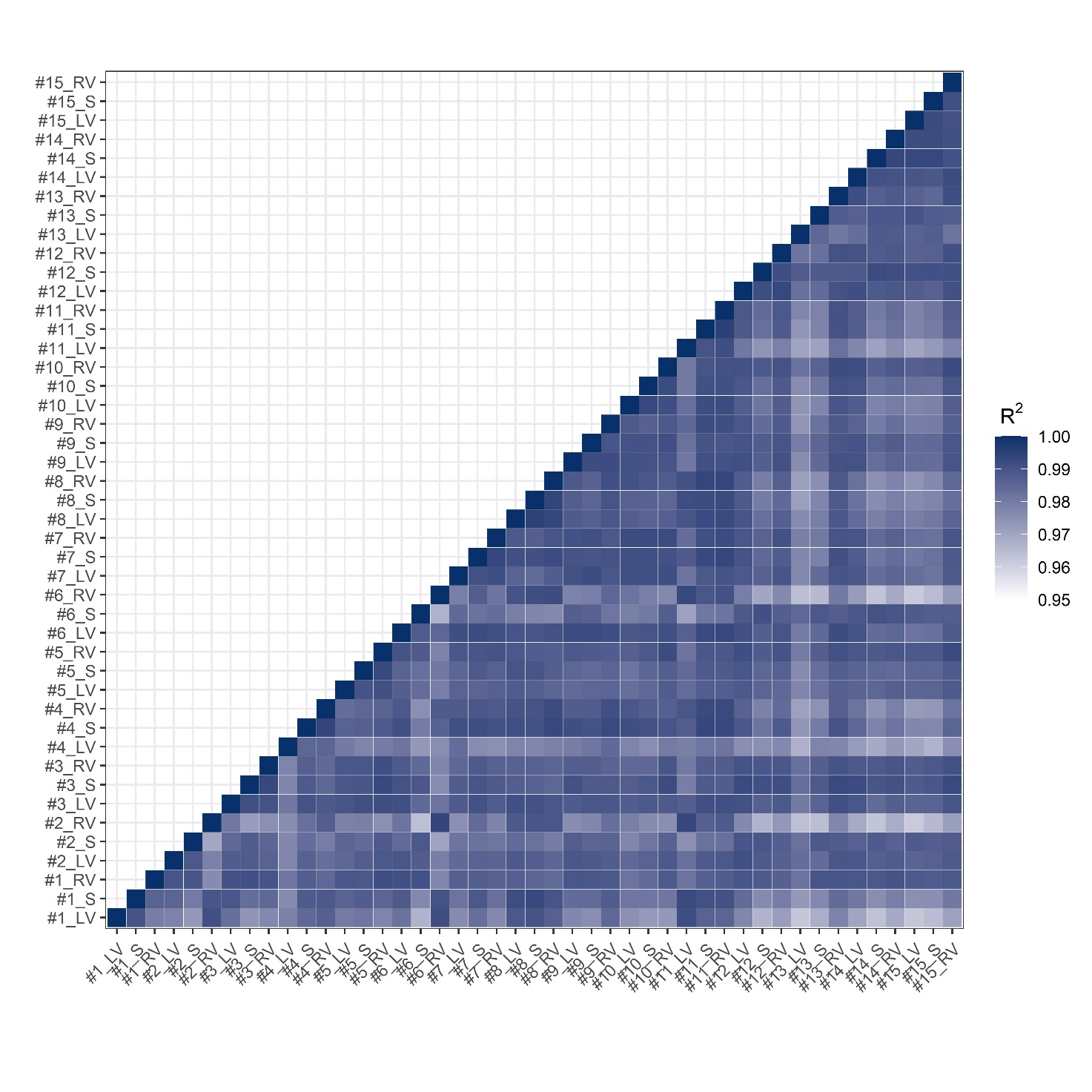

**Correlation of β-values among all samples.** Pairwise comparisons among all samples reported as the squared Pearson's correlation coefficients (R^2^).

#### Supplementary Figure 4

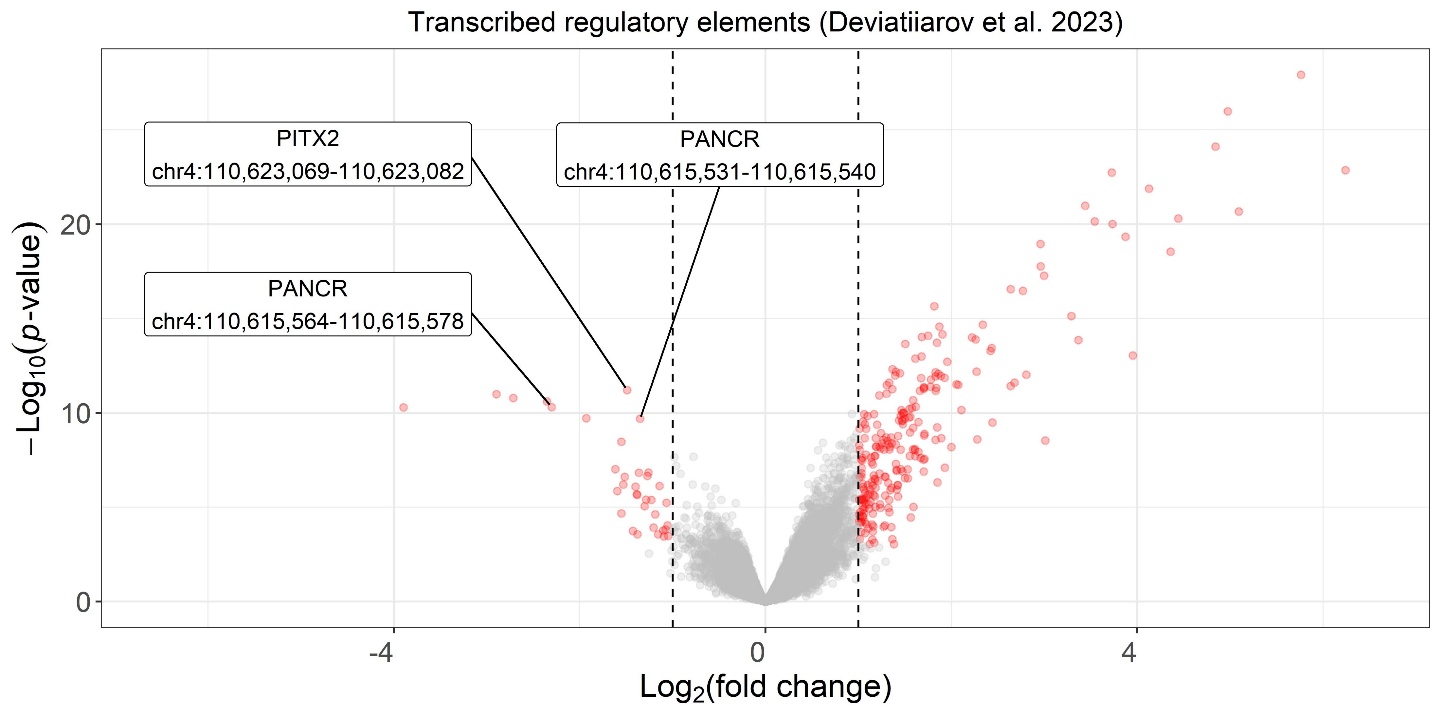

**Volcano plot of transcribed regulatory elements between the left and right side of the heart.** Differential activity of transcribed regulatory elements was examined based on a previous study using cap analysis of gene expression of human hearts (Deviatiiarov et al. 2023). Negative fold change values correspond to the transcribed regulatory elements that are more active in the left than the right side of the heart. Text labels were positioned with the *ggrepel* R package (version 0.9.6).

#### Supplementary Figure 5

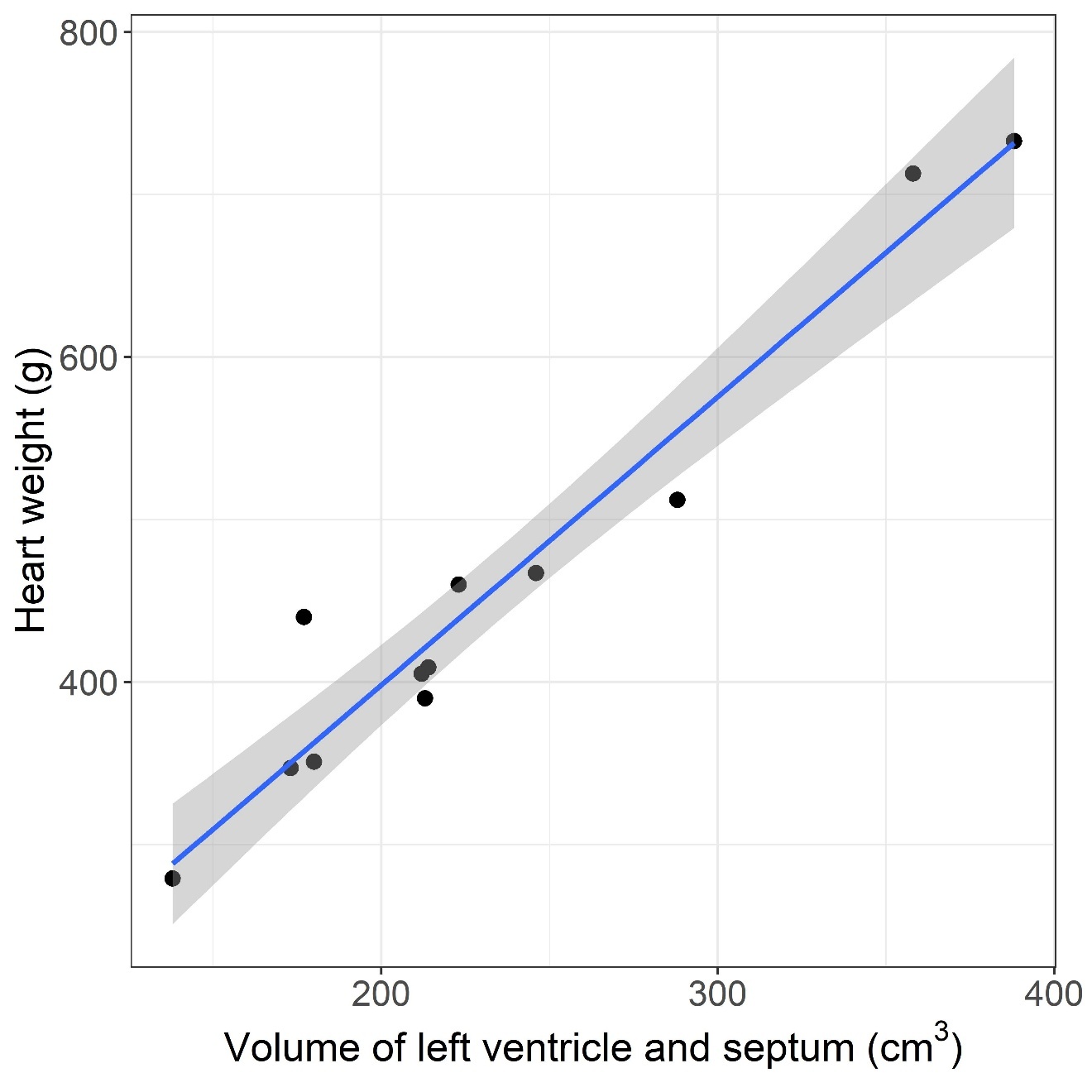

**Autopsy heart weight versus volume of left ventricle and septum estimated from postmortem CT scans**. The regression line from *geom_smooth(method = “lm”)* from the *ggplot* R package is visualised (blue) together with the 95% confidence interval (grey).
